# Astrocytic Synapse Engulfment Is Differentially Controlled by *APOE* Genotype

**DOI:** 10.64898/2026.04.29.26351484

**Authors:** Sowmya Sekizar, Kristjan Holt, Sharon Meyers, Declan King, Jane Tulloch, Rosemary J. Jackson, Tara L. Spires-Jones

**Affiliations:** Institute for Neuroscience and Cardiovascular Research, The University of Edinburgh, UK, EH8 9JZ; UK Dementia Research Institute, University of Edinburgh and University College London, UK; Department of Neuroscience, University of Dundee, UK

## Abstract

*APOE* gene variants encoding the apolipoprotein E (ApoE) protein are strong genetic modifiers of risk of Alzheimer’s disease (AD) with the *APOE* ε4 allele (*APOE4)* associated with substantially increased disease risk, *APOE* ε2 allele (*APOE2*) associated with decreased risk and *APOE* ε3 allele (*APOE3*) considered neutral. Recently the Christchurch variant of *APOE3* (*APOE3Ch*) has been shown to protect people from familial AD. Despite this strong evidence for *APOE* mediating AD risk, the exact biological mechanisms through which *APOE* influences pathogenesis remain unknown. Our previous work implicates *APOE* in synapse degeneration in AD with exacerbated plaque-associated synapse loss, increased accumulation of amyloid beta in synapses, and increased ingestion of synapses by glia around plaques observed in *APOE4* carriers. Here we used a cell culture system to test the hypothesis that *APOE* isoforms would differentially regulate phagocytosis of synapses isolated from human post-mortem AD brain tissue. Humanized *APOE* knock-in astrocyte cell lines exhibited isoform-dependent differences in phagocytic activity with *APOE2* < *APOE3* < *APOE4* as would be expected if *APOE* genotype mediated risk at least in part through synapse phagocytosis. Interestingly, astrocytes with the protective *APOE3Ch* allele phagocytosed synapses similarly to *APOE4* astrocytes indicating this variant does not likely protect from AD by reducing astrocyte phagocytosis of synapses. These findings indicate that *APOE* isoforms differentially regulate astrocytic engulfment of AD-associated synaptic material. These isoform-specific effects are not explained by differences in phosphatidylserine recognition, suggesting the involvement of additional mechanisms underlying ApoE-dependent modulation of astrocyte function.

**Significance Statement:** Sekizar and colleagues tested whether *APOE*, the most important genetic risk factor for Alzheimer’s disease, could affect risk by influencing the ability of astrocytes to phagocytose synapses. Using astrocyte cell lines exposed to synapses isolated from Alzheimer’s disease brain tissue, they demonstrate that different *APOE* isoforms differentially modulate astrocyte-mediated synapse phagocytosis. These results will inform future work to develop therapies that aim to preserve synapses in Alzheimer’s disease.

## Introduction

Alzheimer’s disease (AD) is characterized by progressive cognitive decline, accompanied by extracellular amyloid-β (Aβ) plaques, intracellular tau neurofibrillary tangles, neuroinflammation, and profound neurodegeneration (Braak and Braak, 1995; Thal et al., 2002). Synapse loss is the strongest pathological correlate of cognitive impairment (DeKosky and Scheff, 1990; Terry et al., 1991), preceding overt neuronal death and closely tracking disease severity. Soluble forms of Aβ and tau are found in synapses in human disease brain where they are associated with synapse loss, and they can contribute directly to synapse degeneration in model systems (Colom-Cadena et al., 2023; King et al., 2022; O’Keeffe et al., 2025; Pickett et al., 2019; Tzioras et al., 2023c). Increasing evidence indicates that dysregulated glial-mediated synaptic pruning also contributes to synaptic vulnerability in AD (Beckman et al., 2019; Bie et al., 2019; Dejanovic et al., 2018a, 2018b; Hong et al., 2016; Spurrier et al., 2022). Importantly, preventing synapse loss by blocking synaptic engulfment in mouse models with plaque deposition mitigates cognitive deficits, highlighting the therapeutic potential of targeting glial-mediated synapse clearance (Bie et al., 2019; Hong et al., 2016; Q. Shi et al., 2017; Tzioras et al., 2023a)

The apolipoprotein E (*APOE*) gene encodes a lipoprotein that functions as the primary carrier of cholesterol in the central nervous system, facilitating lipid redistribution from astrocytes to neurons for synaptic repair and maintenance (Liao et al., 2017; Vance and Hayashi, 2010). ApoE is mainly produced by astrocytes and supports membrane repair, synapse formation, and neuronal function. It also helps regulate inflammation, maintain blood–brain barrier integrity, and clear toxic proteins. In *Apoe* knockout mice, loss of ApoE disrupts lipid delivery and neuronal maintenance, leading to increased neuroinflammation, vascular dysfunction, and age-related cognitive impairment (Getz and Reardon, 2012; Piedrahita et al., 1992).

*APOE4* is the strongest genetic risk factor for late-onset AD (Corder et al., 1993). The three common human isoforms—ApoE2, ApoE3, and ApoE4—differ by single amino acid substitutions but exert markedly distinct effects on disease risk (Teter, 2004). *APOE4* significantly increases AD susceptibility and is associated with an earlier age of onset (Corder et al., 1993), whereas *APOE2* confers relative protection and *APOE3* is considered risk-neutral (Corder et al., 1994). Rare variants, such as the Christchurch mutation (*APOE3Ch*), have been associated with delayed onset of cognitive impairment despite increased amyloid burden, compared with individuals without the *APOE3Ch* variant (Arboleda-Velasquez et al., 2019a; Quiroz et al., 2024).

There is strong evidence from human tissue and multiple model systems that ApoE is involved in pathological protein aggregation and clearance (Huynh et al., 2017; Lin et al., 2018; Liu et al., 2013; Nixon, 2017; Y. Shi et al., 2017). In addition to these roles, there is accumulating evidence that ApoE is involved in synapse degeneration in AD. We observed that ApoE and amyloid beta co-localize within a subset of synapses in human AD brain (Koffie et al., 2012a).Synaptic amyloid beta accumulation associates with synapse loss around plaques and is exacerbated in *APOE4* carriers (Jackson et al., 2019; Koffie et al., 2009). In model systems, amyloid beta is directly synaptotoxic (Borlikova et al., 2013; Koffie et al., 2009; Shankar et al., 2007; Spires-Jones et al., 2009) and ApoE4 increases synaptic localization of amyloid beta (Koffie et al., 2012b). These data imply that one pathological role of *APOE4* is exacerbating synapse loss through directly bringing toxic amyloid beta to synapses.

Growing data also implicate *APOE* in neuroinflammation (Tzioras et al., 2019). *APOE* is upregulated in disease-associated microglia around plaques in both mouse model systems and human post-mortem tissue (Ulrich et al., 2018; Wang et al., 2024), which could contribute to both clearance of amyloid beta and microglial-mediated synaptic engulfment observed in animal models and human tissue(Hong et al., 2016; Tzioras et al., 2023b).Astrocytes are the primary source of ApoE in the CNS and play essential roles in synapse formation, maintenance, and elimination. In both development and disease, astrocytes contribute to synaptic remodelling through phagocytosis, acting in concert with or independently of microglia (Chung et al., 2013; Lee et al., 2021; Vainchtein et al., 2018). In AD, astrocytes adopt reactive phenotypes and exhibit altered lipid metabolism, inflammatory signalling, and lysosomal function—processes that may be modulated by *APOE* genotype(Liddelow et al., 2017). Although microglial complement-mediated synaptic pruning has been widely studied in AD (Bie et al., 2019; Hong et al., 2016), less is known about how astrocytic ApoE regulates the engulfment of synaptic material(Mallach et al., 2024), particularly in response to AD-associated pathological cues. Previous work from our lab demonstrated that post-mortem human tissue from AD brains shows increased synaptic protein localized within microglia and astrocytes compared with controls, with this ingestion further elevated in *APOE4* carriers. Opsonins such as MFGE8 bind to phosphatidylserine (PS) “eat-me” signals exposed on synaptic membranes, facilitating recognition by integrin receptors on astrocytes. Blocking MFGE8 or inhibiting integrin receptor function significantly reduces phagocytosis in cultured human and mouse astrocytes challenged with synaptoneurosomes derived from AD cases (Tzioras et al., 2023a).

The phagocytic roles of astrocytes have gained increasing attention following recent findings demonstrating that mice lacking microglia exhibit largely normal synaptic remodelling, maturation, and behaviour, suggesting that astrocytes can independently contribute to synapse clearance(Li et al., 2024; O’Keeffe et al., 2025). Astrocytes can engulf synapses through phagocytic receptors such as MEGF10 and MERTK(Chung et al., 2013). Together, these findings highlight a receptor- and opsonin-mediated mechanism by which astrocytes recognize and clear synaptic material. Whether ApoE isoforms differentially modulate astrocytic responses to these signals remains unknown.

Here we test the hypothesis that *APOE* isoforms differentially regulate astrocyte phagocytosis of synapses using astrocyte cell lines exposed to synaptic enriched fractions isolated from post-mortem human AD brain tissue.

## Results

### *APOE* Isoforms differentially regulate synapse phagocytosis

Humanized *APOE* knock-in (KI) astrocyte lines were immunolabelled for glial fibrillary acidic protein (GFAP), confirming astrocytic identity across all genotypes (**Fig. 1A**). Phagocytic activity was assessed using pHrodo-labelled human AD synaptoneurosomes applied to cells and live-cell imaging over 48 hours. Representative images at 48 hours are shown in **Fig. 1B**. As pHrodo fluorescence is only emitted upon internalisation into acidic lysosomal compartments, the fluorescent signal reflects phagocytic activity. The area of internalised pHrodo signal was quantified and normalised to total cell area, derived from corresponding brightfield images. This ratio was plotted over time (**Fig. 1C**). Quantification of internalised pHrodo signal, normalised to cell area, revealed a time-dependent increase in phagocytosis across all genotypes (**Fig. 1C**, ANOVA after linear mixed effects model *APOE* genotype *time + (1|plate/well) on Tukey transformed data, effect of time F(24,1111) = 142.20, p < 2 × 10^−16^). Uptake also differed significantly by *APOE* genotype (F(4,1111) = 53.39, p < 2 × 10^−16^)). *APOE2* astrocytes had significantly reduced phagocytic activity compared to all other genotypes (Tukey-corrected post-hoc pairwise comparisons p<0.05 for *APOE2* vs each other genotype). *APOE4* astrocytes had increased synapse phagocytosis compared to *APOE2* (estimate = 0.11, t = −11.45, p < 0.0001) and *APOE3* (estimate = 0.05, t = −4.953, p < 0.0001), but were not significantly different from *APOEKO* or *APOECh* cells. *APOE3* astrocytes also showed significantly lower uptake compared to *APOEKO* cells (estimate = −0.03, t = −3.29, p = 0.01) and *APOEch* cells (estimate = −0.06, t = −6.55, p < 0.0001).

**Figure 1:**
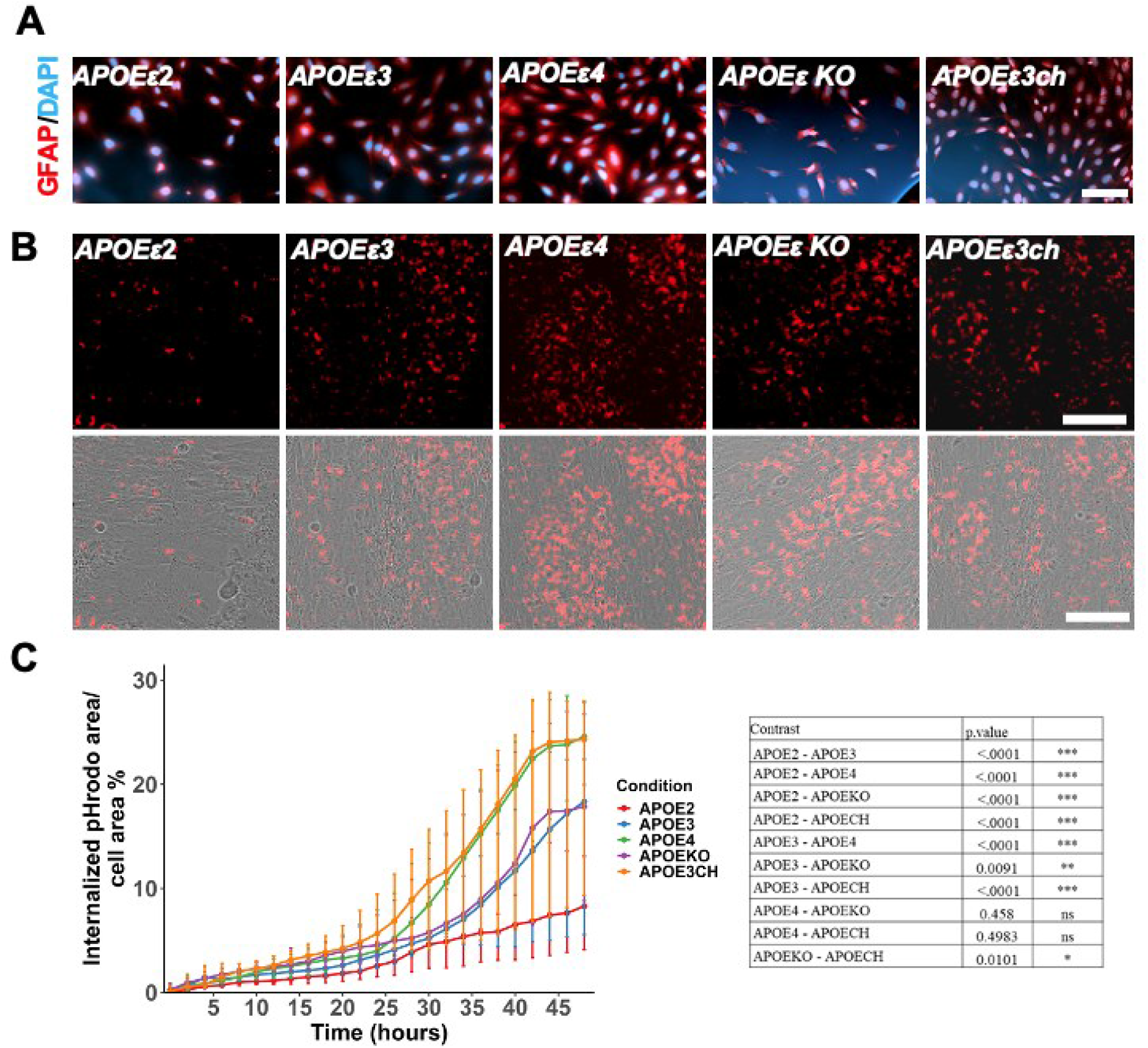
Differential phagocytosis of AD synapses by astrocytes depending on *APOE* genotype. Immunocytochemical characterization of humanized APOE knock-in (KI) astrocytes expressing *APOE2, APOE3, APOE4, APOE3Ch* or with a knockout of mouse *Apoe* (*APOEKO*) were stained with astrocyte marker GFAP (red) and DAPI (blue) confirming lineage identity (**A**). Representative images of the Live-imaging assay of APOEKI astrocytes shows astrocytes phagocytose pHrodo-Red-labelled human synaptoneurosomes from Alzheimer’s disease (AD) brains after treatment (**B**). Quantification of phagocytosis of pHrodo labelled synapses shows increases in ingestion over time with a significant difference between genotypes (**C**). Significant post-hoc comparisons between genotypes are indicated with brackets on the legend. Scale bars represent 100 μm in panel A and 50 μm in panel B.

These data demonstrate *APOE* isoform–dependent regulation of astrocyte-mediated engulfment of AD-derived synaptoneurosomes in our model system. *APOE4* and *APOE3ch* exhibited the highest phagocytic capacity, *APOE3* and *APOEKO* showed intermediate levels, and *APOE2* displayed a markedly attenuated response. The absence of a significant genotype-by-time interaction indicates that isoforms differ primarily in overall magnitude of uptake rather than temporal dynamics.

### Phosphatidylserine-Dependent Uptake Is Not Differentially Regulated by *APOE* Isoforms

Externalized phosphatidylserine (PS) functions as a canonical “eat-me” signal during synaptic pruning and phagocytosis, frequently requiring opsonins for receptor-mediated recognition. To determine whether the *APOE* isoform–dependent differences observed in synaptoneurosome phagocytosis were attributable to altered PS recognition, we generated PS-containing liposomes (formulated according to established ratios in (Scott-Hewitt et al., 2020) and labelled them with the lipophilic dye DiO (PS-DiO). These particles contained PS without synaptic proteins or other neuronal components, enabling selective assessment of PS-driven uptake **(Fig.2A)**

**Figure 2.**
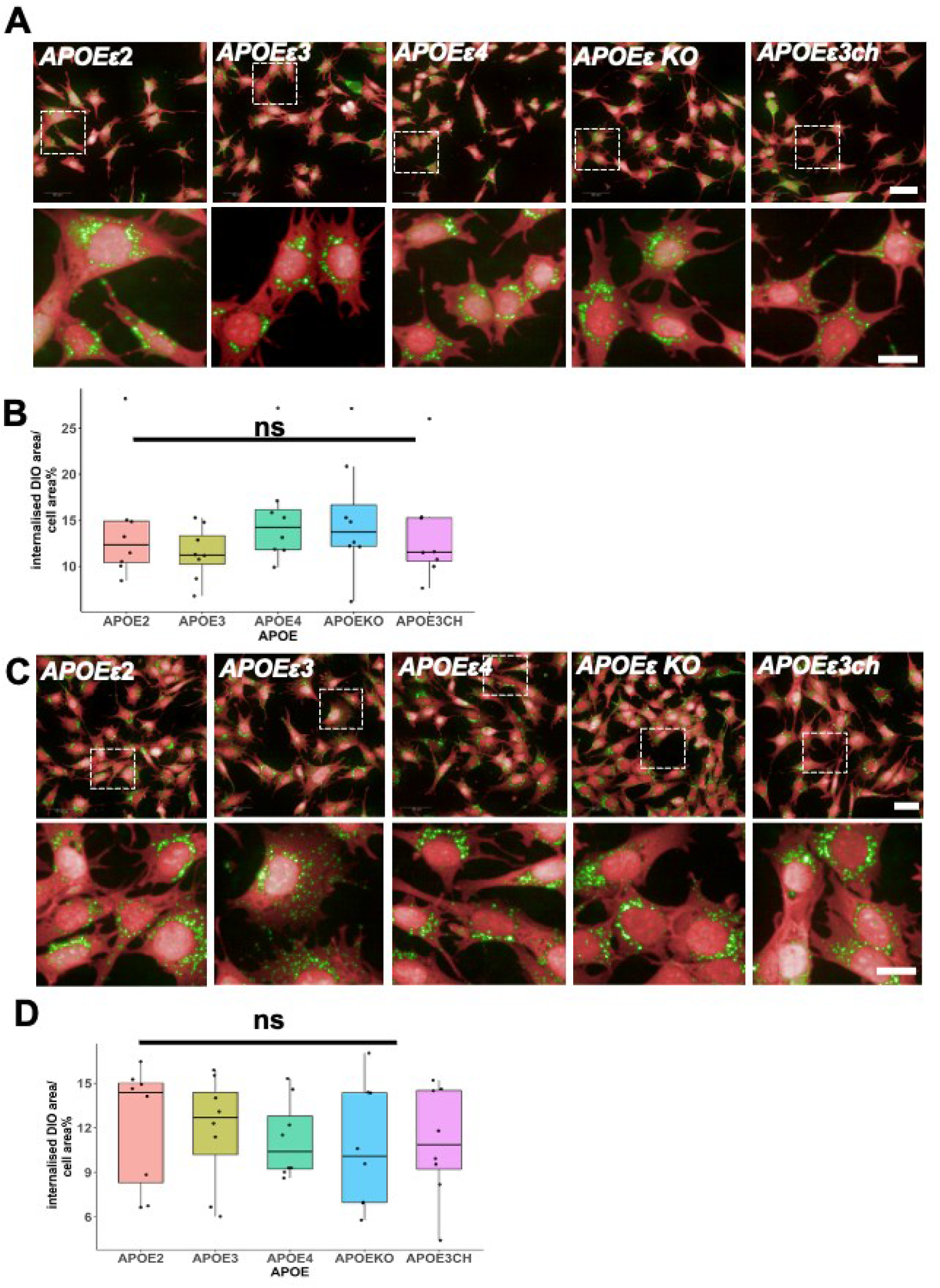
Phosphatidylserine uptake is not differentially regulated by *APOE* isoforms. Astrocytes with different *APOE* genotypes were treated with PS-DiO (green) for 48 hours then rinsed fixed and stained with CellMask (red) (**A**). *APOEKO* astrocytes were treated with PS-DiO that had been pre-incubated with conditioned media from astrocytes of different *APOE* genotypes (B). Quantification of internalized PS-DiO showed no difference between APOE genotypes of astrocytes (**C**, Type III ANOVA on Tukey-transformed data showed no significant effect of APOE genotype (F(4,28) = 1.95, p = 0.129) or conditioned media (D, F(4,28) = 0.665, p = 0.6216). Scale bars represent 100μm (main panels) 20 μm (insets). Individual data points represent culture replicates (n=7 per condition).

PS-DiO was first applied directly to astrocytes of different *APOE* genotypes for 48 hours. Cells were fixed and stained with CellMask to delineate cytoplasmic boundaries, allowing accurate quantification of internalised fluorescence, with nuclei identified by DAPI (**Fig. 2A**). To assess whether secreted APOE isoform–specific factors influence PS handling, conditioned media (CM) from each genotype were pre-incubated with PS-DiO prior to application to *APOEKO* astrocytes. Following 48 hours, uptake was quantified as above (**Fig 2B**). Quantitative analysis revealed no significant effect of astrocyte *APOE* genotype on PS-DiO uptake (**Fig 2C**, Type III ANOVA: F(4, 28) = 1.95, p = 0.129) and no significant effect of conditioned media *APOE* genotype on PS-DiO uptake (**Fig 2D**, F(4, 28) = 0.665, p = 0.622).

Together, these data demonstrate that neither intrinsic *APOE* isoform expression nor *APOE* isoform–specific secreted factors alter the uptake of PS-containing liposomes. These findings indicate that differential phosphatidylserine recognition alone does not account for the isoform-dependent differences observed in AD-derived synaptoneurosome phagocytosis.

## Discussion

Our findings demonstrate that humanized ApoE isoforms differentially regulate astrocyte-mediated phagocytosis of AD-derived synaptoneurosomes in immortalized astrocyte cell culture. Consistent with the hypothesis that ApoE isoforms influence AD risk in part through mediating synaptic phagocytosis by astrocytes, *APOE2* astrocytes exhibited the lowest levels of synapse phagocytosis with higher levels in *APOE3* and higher still in *APOE4* astrocytes. Surprisingly, *APOE3Ch* astrocytes ingested similar amounts of synapses to *APOE4* astrocytes, indicating that the protective effect of *APOE3Ch* in familial AD is not likely mediated by affecting astrocyte phagocytosis of synapses.

Previous work by (Chung et al., 2016), demonstrated that ApoE isoforms differentially regulate astrocyte-mediated synapse pruning in vivo, with ApoE2 enhancing and ApoE4 reducing physiological synaptic turnover. *APOE* genotype also influenced age-dependent accumulation of complement component C1q at synapses, suggesting that *APOE2* promotes efficient clearance of senescent synapses, whereas *APOE4* is associated with reduced astrocytic phagocytic capacity and greater accumulation of synaptic debris. In contrast, our study utilized synaptoneurosomes derived from Alzheimer’s disease (AD) patient brain, rather than synaptosomes from healthy mouse tissue. These synapses from end-stage AD contain Aβ, tau, complement proteins, and other opsonins (Hesse et al., 2019). The differing engulfment observed in *APOE* astrocytes in our system may therefore reflect a context-dependent response to pathologically altered synaptic material rather than baseline synaptic maintenance.

The APOE Christchurch (*APOE3Ch*) variant has recently been associated with protection against Alzheimer’s disease (AD) pathology in people with familial AD *PSEN1* variants (Arboleda-Velasquez et al., 2019b). In mouse models of tauopathy, *APOE3Ch* rescues synapse loss, and microglia expressing the *APOE3Ch* variant engulf more tau while limiting tau seeding and spread. Additionally, myeloid cells carrying *APOE3Ch* have been shown to exhibit enhanced tau phagocytosis (Chen et al., 2024). However, the role of *APOE3Ch* in astrocyte-mediated synapse phagocytosis has not been previously characterized, and little is known about its impact on astrocytic engagement with synapses. In our astrocyte model, *APOE3Ch* cells demonstrated greater phagocytic activity than *APOE2*, suggesting that the protective effects of *APOE3Ch* may not arise from reduced synapse phagocytosis.

Our phosphatidylserine (PS)-based assays indicate that the *APOE* isoform-dependent differences in synapse phagocytosis are not driven by altered PS recognition alone. This suggests that other mechanisms—such as complement-mediated opsonisation, receptor signalling, or lysosomal processing—are more likely to underlie the observed effects.

Several limitations should be considered. The use of a simplified *in vitro* system does not fully capture the complexity of the *in vivo* brain environment. Additionally, the requirement for serum to maintain cell viability may have induced an activated astrocyte state, potentially influencing phagocytic behaviour and amplifying genotype-dependent differences. Future work should focus on defining the mechanisms and physiological relevance of *APOE*-dependent synaptic engulfment. First, experiments in serum-free or chemically defined conditions, as well as co-culture systems incorporating neurons and microglia, will be essential to better model the in vivo environment. Second, it will be critical to determine whether ApoE isoforms drive selective clearance of dysfunctional synapses—such as those enriched in Aβ or tau—or promote indiscriminate synaptic loss. This could be addressed using labelled healthy versus diseased synaptic material or live imaging approaches to track synapse fate. Third, the role of opsonins should be investigated in detail. Complement proteins (e.g., C1q, C3) are key mediators of synaptic tagging, and ApoE isoforms may differentially regulate their deposition, receptor engagement (e.g., CR3, MEGF10, MERTK), or downstream signalling pathways. Dissecting these interactions will clarify how *APOE* integrates with established synaptic pruning mechanisms. Fourth, intracellular processing pathways—including endolysosomal trafficking and degradation efficiency—should be examined to determine whether ApoE isoforms influence not only uptake but also cargo handling. Finally, *in vivo* validation using humanized *APOE* models will be essential to establish the relevance of these findings to disease progression. In particular, assessing synapse density, complement deposition, and astrocyte reactivity across *APOE* genotypes in AD models will help determine whether enhanced astrocytic phagocytosis contributes causally to synapse loss and cognitive decline.

Together, this work supports a model in which ApoE isoforms shape astrocyte-mediated synaptic clearance in a context-dependent manner, with important implications for synapse vulnerability and neurodegeneration in Alzheimer’s disease.

## Material and Methods

### Immortalized *APOE* astrocyte cell lines

*Apoe* knockout mouse astrocytes (Morikawa et al., 2005) were transduced with lentivirus according to the Lentivirus Transduction Protocol by Vectorbuilder. HiBiT-tagged ε2, ε3, ε3 Christchurch, and ε4 lentivirus was added to the cells, adapted to a Multiplicity of Infection (MOI) of 5 along with 6ug polybrene per well. DMEM was used for a media change after 20h and subsequent cell maintenance. Successfully transduced astrocytes were selected in a 2-week incubation period with 8ul Hygromycin B (Invitrogen, #10687010) per well. (ScienCell #1801). Successful transduction was measured following instructions of the Nano-Glo HiBiT Lytic Detection System measured in the Perkin-Elmer Victor 3V luminescence plate reader. Single clones were selected and qPCR for *APOE, Gfap*, and *Actin* were run to determine expression of APOE. RNA was extracted using the RNeasy Mini Kit (Qiagen) and eluted in 10 μl of nuclease-free water. RNA was retrotranscribed to complementary DNA using the QuantiTect Reverse Transcription kit (Qiagen). RT–qPCR was conducted with RT^2^ SYBR Green Mastermix fluorescent dye (Qiagen) and QuantiTect primers (Qiagen). Clones with similar *APOE* expression were used for this experiment to ensure that the expression of APOE did not differ between isoforms.(supplementary data, figure 2)

*APOE* cell lines were cultured in DMEM F-12 (Gibco,21041-025), pen strep 1% (Gibco, 15140-122), FBS 10% (A5256701). Frozen cells were thawed and plated in T75 flasks, after two passage the cells were treated with geneticin (Gibco,10131-035) to select for KI cells. To plate cells, the medium was removed, cells dislodged using trypsin 0.05%(Invitrogen,25200056). Add 5ml of medium to neutralize trypsin, collect cells suspension spin at 2500rpm and 2.5 min. The cells were resuspended in medium and plated at a density at 2000-3000 cells per 96 well plate (Greiner,655180). The plates were not coated with PDL. The cells were treated with DIO-PS 2 days after plating.

### Immunofluorescence Staining

Cells were fixed in 4% paraformaldehyde (PFA) and rinsed with 0.1 M phosphate-buffered saline (PBS). Following fixation, cells were permeabilized and blocked in a solution containing 10% fetal bovine serum (FBS) and 0.1% Triton X-100 for 1 hour at room temperature. Cells were then incubated with a primary anti-GFAP antibody (chicken, ab4674) diluted 1:500 in blocking solution for 2 hours at room temperature. After incubation, the primary antibody was removed, and cells were washed thoroughly with 1× PBS. Nuclei were counterstained with DAPI, followed by final washes in PBS prior to imaging.

### Human Brain Tissue and Synaptoneurosome Preparation

Human brain tissue from Alzheimer’s disease was obtained from [UK brain bank network] with appropriate ethical approval.All tissue was provided by the MRC Edinburgh Brain Bank, following all appropriate ethical approval and informed consent of the donor’s pre-mortem and their families’ post-mortem cases were cross-checked neuropathologically and were confirmed to be Braak Stages V-VI. Data about subjects included in the study are found in Table S1. Use of human tissue for post-mortem studies has been reviewed and approved by the Edinburgh Brain Bank ethics committee and the ACCORD medical research ethics committee, AMREC (approval number 15-HV-016; ACCORD is the Academic and Clinical Central Office for Research and Development, a joint office of the University of Edinburgh and NHS Lothian). The Edinburgh Brain Bank is a Medical Research Council funded facility with research ethics committee (REC) approval (11/ES/0022)

### Synaptoneurosome preparation

Preparation of synaptoneurosomes was performed as described previously(Tai et al., 2012). Snap-frozen human tissue of 300-500 mg from BA38 and BA20/21 (temporal cortex) was homogenised using a Dounce homogeniser with 1mL of a protease inhibitor buffer, termed here Buffer A. Buffer A consists of 25 mM HEPES, 120 mM NaCl, 5 mM KCl, 1 mM MgCl2, 2 mM CaCl2, protease inhibitors (Merck, 11836170001) and phosphatase inhibitors (Merck, 524629-1SET) made up in sterile water, and was prepared fresh each day. The Dounce homogeniser and Buffer A were kept ice-cold throughout the procedure to avoid further degradation. We allowed 10 passes for full homogenization but minimise cellular disruption that would lead to an impure final fraction. Once homogenised, the homogenate was aspirated in a 1mL syringe and passed through an 80-micron filter (Merck, NY8002500) to remove debris and yielded the total homogenate (TH). The filter was pre-washed with 1mL of Buffer A to maximise yield. A sample of the TH was snap-frozen on dry ice for Western blot analysis, and the rest was split in half for Western blot analysis and the phagocytosis assays. A subsequent filtration took place using a 5-micron filter (Merck, SLSV025LS) followed by centrifugation at 1000 x g for 7 min to yield the synaptoneurosome (SN) pellet. In this step, extra care was taken to slowly pass the homogenate through the filter in order to prevent the filter breaking. The filter was pre-washed with 1mL of Buffer A to maximise yield. The pellet was washed with Buffer A and pelleted down again to ensure purity. Pellets were snap frozen on dry ice and stored at −80°C for long-term storage.A total 11 samples from AD donors and 11 non-dementia control donors were used(supplementary data, table 1).

**Table 1.**

| Age- in<br>their | APOE<br>genotype | Braak<br>Stage | BBN |
| --- | --- | --- | --- |
| 80's | 4/4 | VI | BBN0011.265 |
| 70's | 4/4 | VI | 15810 |
| 70's | 3/4 | VI | BBN001.374 |
| 90's | 3/3 | VI | BBN001.37667 |
| 60's | 3/3 | VI | BBN001.35182 |
| 90's | 3/4 | VI | BBN001.29135 |
| 70's | 3/4 | VI | BBN_24526 |
| 90's | 3/4 | VI | BBN_24668 |
| 70's | 3/4 | VI | BBN001.37702 |
| 70's | 3/4 | VI | BBN001.35096 |
| 76's | 3/4 | VI | BBN001.37808 |

### Protein extraction

For protein extraction, samples we diluted 5-fold with Tris-HCl buffer pH 7.6 (100 mM Tris-HCl, 4% SDS, and Protease inhibitor cocktail EDTA-free [Thermo Fisher Scientific, 78447]), followed by centrifuging for 25 min at 13,3000 RPM at 4°C. Then, the supernatant was collected in fresh tubes and heated at 70°C for 10 min. Samples were kept at −80°C for long-term storage.

### Micro BCA

Micro BCA Protein Assay kit (Thermo Fisher Scientific, 23235) was used to quantify protein levels, following manufacturer’s instructions. Briefly, 1 μL of sample was added in 1 mL of working solution and heated at 60°C for 1 h. Working solution was made fresh right before required, consisting of 50% Buffer A, 48% Buffer B and 2% Buffer C, all provided in kit. Albumin was provided in the kit for determining a standard curve of the following concentration: 2 μg/mL, 4 μg/mL, 6 μg/mL, 8 μg/mL, 10 μg/mL,20 μg/mL and 40 μg/mL. Absorbance values were obtained using spectrophotometry at 562 nm with working solution used as the blank value to calibrate the machine.

### Western blot

After protein extraction, each sample was made-up to 20 μg of protein as calculated by the micro BCA (protein extract diluted in de-ionised water) and diluted in half with Laemmli buffer (2x stock) (S3401-10VL). In each well, 15 μL of sample were loaded in 4–12% Bis-Tris gels (ThermoFisher Scientific, NP0323BOX). Each gel was run with 5 μL of molecular weight marker (Licor, 928–40000) in the first well. Gels were washed with de-ionised water and diluted NuPAGE buffer (ThermoFisher Scientific, NP0002) (20x stock) to remove bits of broken gel and residual acrylamide. Western blot chambers were filled with diluted NuPAGE buffer (600 mL per chamber), making sure each chamber compartment was locked securely and there was no leaking. Gels were run at 80 V for 5 min, 100 V for 1.5 h and 120 V for 30 min. Then, using a scraper tool, gels were cracked open from their plastic casing and soaked in 20% ethanol for 10 min prior to transferring using the iBlot 2 Dry Blotting System (IB21001). Pre-packed transfer stacks containing a PVDF membrane (ThermoFisher Scientific, IB24002) were assembled as per manufacturers recommendations, and samples were transferred for 8.5 min at 25 V. After transferring, the PVDF membranes were stained for 5 min with REVERT protein stain (RT). Wash briefly in LICOR Wash Buffer. Take membrane to LICOR Imager with tweezers. Exposure Auto 700 nm for total protein signal. Wash membrane in Reversal REVERT buffer for 5 mins, rocking (RT)Briefly wash membrane in PBS-T and were blocked using Odyssey blocking buffer (Licor, 927–40000) for 30 min, following overnight incubation with primary antibodies made up in Odyssey block with 0.1% Tween 20, at 4°C with gentle shaking. The following primary antibodies were used: Synaptophysin (mouse, Abcam ab8049, 1:1000), PSD-95 (rabbit, Cell signalling D27E11, 1:1000) and Histone (rabbit, Abcam ab1791, 1:1000).The next day, membranes were washed 6 times with PBS-0.1% Tween 20, and incubated with the following LI-COR secondary antibodies for 30 min: IRDye 680RD Donkey anti-Mouse IgG, highly cross-adsorbed (LI-COR, 925–68072, 1:5000) and IRDye 800CW Donkey anti-Rabbit IgG, highly cross-adsorbed (LI-COR, 925–32213, 1:5000). All gels were imaged using the same exposure times and intensities using an LI-COR Scanner. The images were then uploaded on Image Studio for analysis. For each band, the same size box was used to ensure all samples are measured equally. Each sample was normalized to its corresponding value of total protein.(supplementary data, figure 1)

### Quantitative PCR

Total RNA was extracted using the Qiagen RNeasy Mini Kit according to the manufacturer’s instructions. qPCR was performed using Qiagen kits with gene-specific primers from Qiagen on a real-time PCR system. Gene expression was normalised to actin.

### Phagocytosis Assay

Due to the long image-acquisition times acquired for astrocyte phagocytosis assays, these experiments were performed in an IncuCyte S3 live-imaging system that is housed in a tissue-cultured incubator. Phase images were taken prior to synaptoneurosome addition to normalize ingestion for the confluency of astrocytes per well. Astrocytes were imaged every 2 h with the 20X objective for up to 48 h on phase contrast and red channel to pick up the pHrodo-Red signal. PHrodo-Red area was normalised to cell area.

### Synaptoneurosome and synaptosome labelling with pHrodo Red-SE

First, pHrodo Red-SE (ThermoFisher Scientific, P36600) was diluted with DMSO according to manufacturer’s instructions to reach a concentration of 10 mM. Synaptoneurosome and synaptosome pellets were resuspended in 100 mM sodium carbonate buffer pH 9, adapted from a previously described protocol.28 Synaptoneurosomes were tagged with pHrodo Red-SE based on protein concentration, roughly at 4 mg/mL, as calculated by the micro-BCA, and incubated at room temperature under gentle shaking for 1 h, covered in foil. Samples were centrifuged at 13,000 RPM for 10 min to obtain the labelled synaptoneurosome pellet, followed by 3 rounds of washing with PBS and centrifugation to wash out any unbound dye. The pHrodo-labeled synaptoneurosome pellets were then resuspended in 5% DMSO-PBS and aliquoted for storage at −80°C. Aliquots of pooled AD samples were prepared fresh.

### Liposome’s preparation and assay

Liposomes were prepared using 99% phosphatidyl serine and 1% DIO, mol %. by avanti lipids. According to protocol Scott-Hewitt, 2020. Liposomes were used at 0.01mM. The cells were starved for 1hr with no serum medium and then PS-DIO was added to cell and cells maintained at 37Deg C, 5% CO2 for 48hour, fixed with PFA4% for 10min and rinse in PBS1X and add cell mask (1:200, Thermo fischer,10033484) for 30min and then rinse with PBS1X and stain nuceli with DAPI. Rinse with PBS1X and the plate was imaged on confocal,40 x air objective, Opera Phenix Plus high-content screening system. For condition media experiment, medium from APOE cells were collected, the medium spun down at 12000rpm to remove cell debris and the medium was incubated with PS-DIO and for 2 hrs and this suspension was added to *APOEεKO* cells.

### Quantification and statistical analysis

Statistical analyses were performed in R Studio54 using R version 4.1.2. For data from incucyte and OPERA the data was analyzed using LMM, To investigate differences between groups, linear mixed effects models were used. Statistical analysis was performed using a linear mixed-effects model (lmer, *lme4* package in R):% pHrodo area per cell area ∼ APOE genotype * Time + (1 | plate/well).

Plate was included as a random intercept to account for variability between experimental plates and to account for duplicate measurements from the same genotype on each plate. Each plate was from a different passage of cells. Data are presented as median ± IQR unless otherwise indicated.

## Data Availability

All data produced in the present study are available upon reasonable request to the authors

## Acknowledgements

We would like to thank brain tissue donors and their families for providing tissue donations, without which this work would not be possible. We gratefully acknowledge David Holtzman and Brad Hyman for donating materials and support and Edinburgh Neuroscience and the FENS-Kavli Network of Excellence for facilitating collaborations leading to this work. We gratefully acknowledge the contributions of the Edinburgh Brain and Tissue Bank and Alzheimer’s Scotland Dementia Research Centre for coordinating post-mortem brain tissue donations.

---

**Supplementary figure 1:**
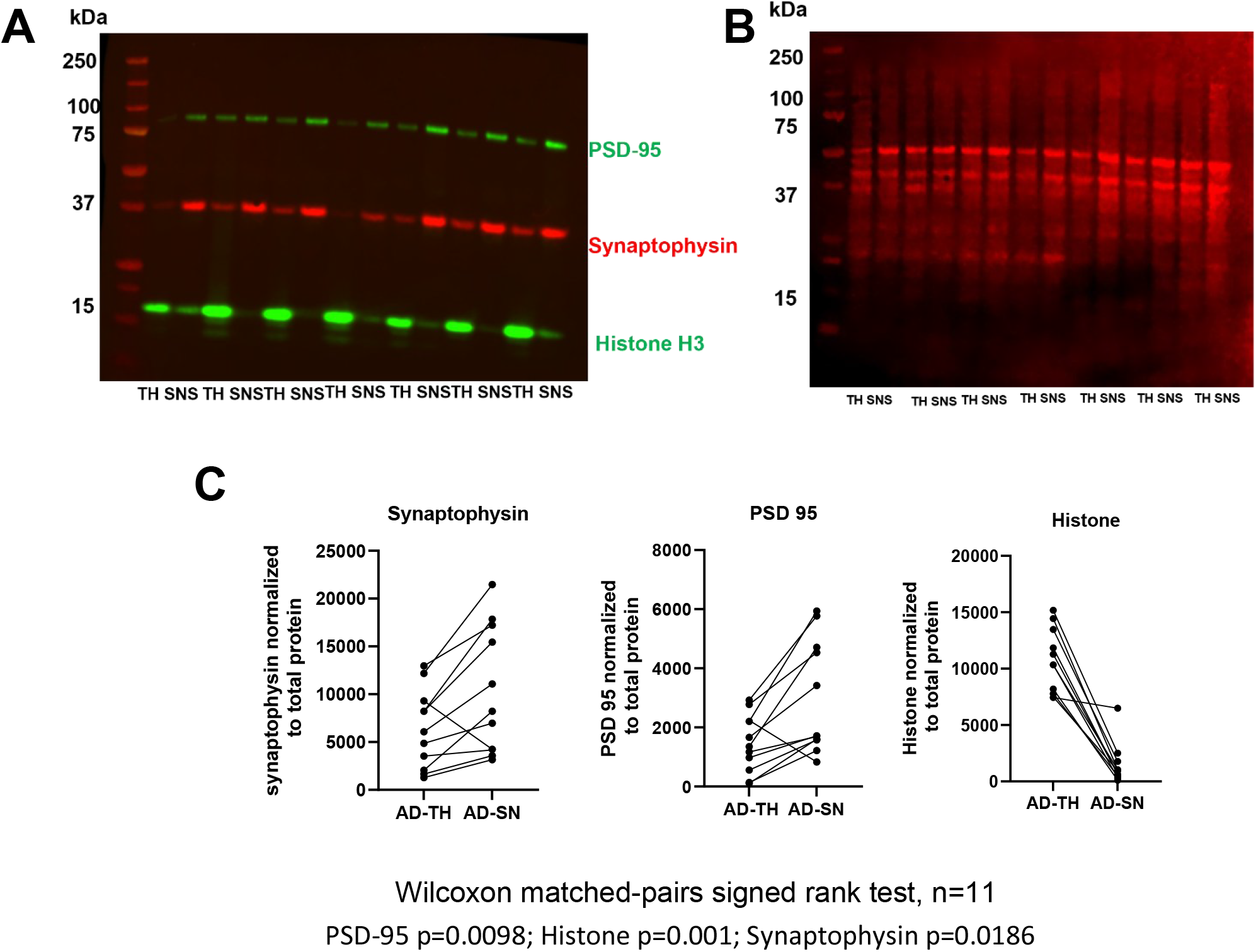
Validation of synaptoneurosome and synaptosome preparations. Image of a full-length Western blot (**A**), indicating whether a sample is from total homogenate (TH) or synaptoneurosomes (SN). Bands were quantified on Licor, and normalised to total protein stain shown in (**B**). Quantification (**C**) shows significantly increased protein levels of the pre- and post-synaptic markers synaptophysin and PSD-95, respectively, and decreased protein levels of histone (H3), indicating exclusion of non-synaptic material (Wilcoxon matched-pairs signed rank test).

**Supplementary figure 2:**
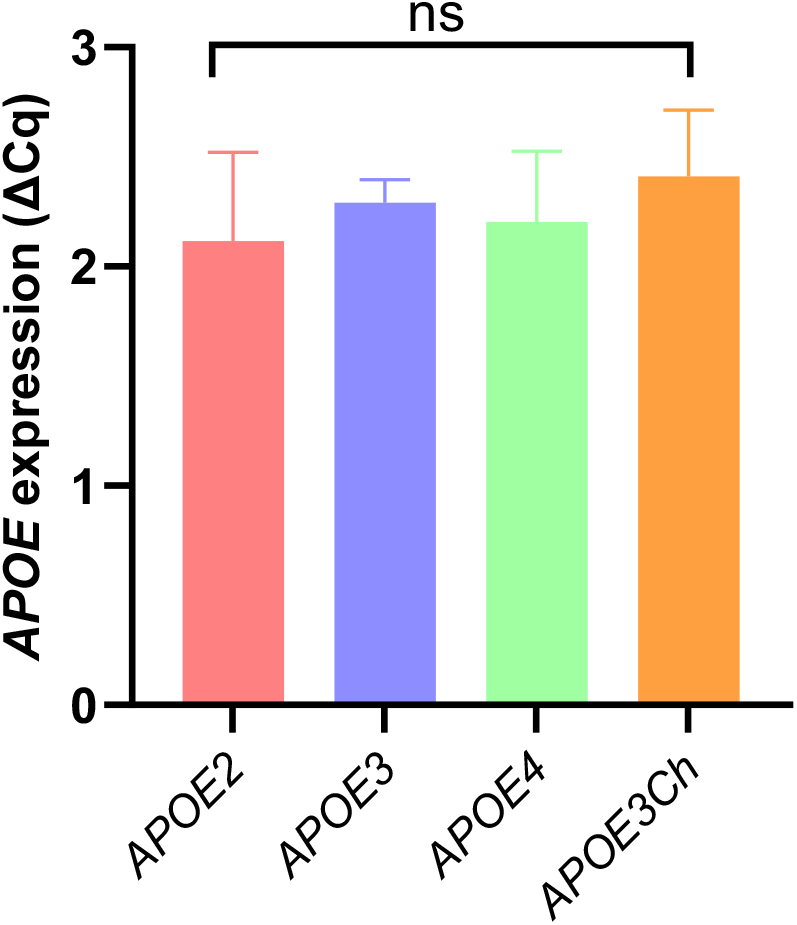
Comparable APOE expression across engineered cell lines normalised to actin. qPCR analysis shows that APOE expression levels are consistent across all APOE-modified cell lines when normalised to the housekeeping gene actin, indicating that humanized APOE insertion does not alter expression levels.

