## Supplementary information for "Astrocytic Synapse Engulfment Is Differentially Controlled by *APOE* Genotype"

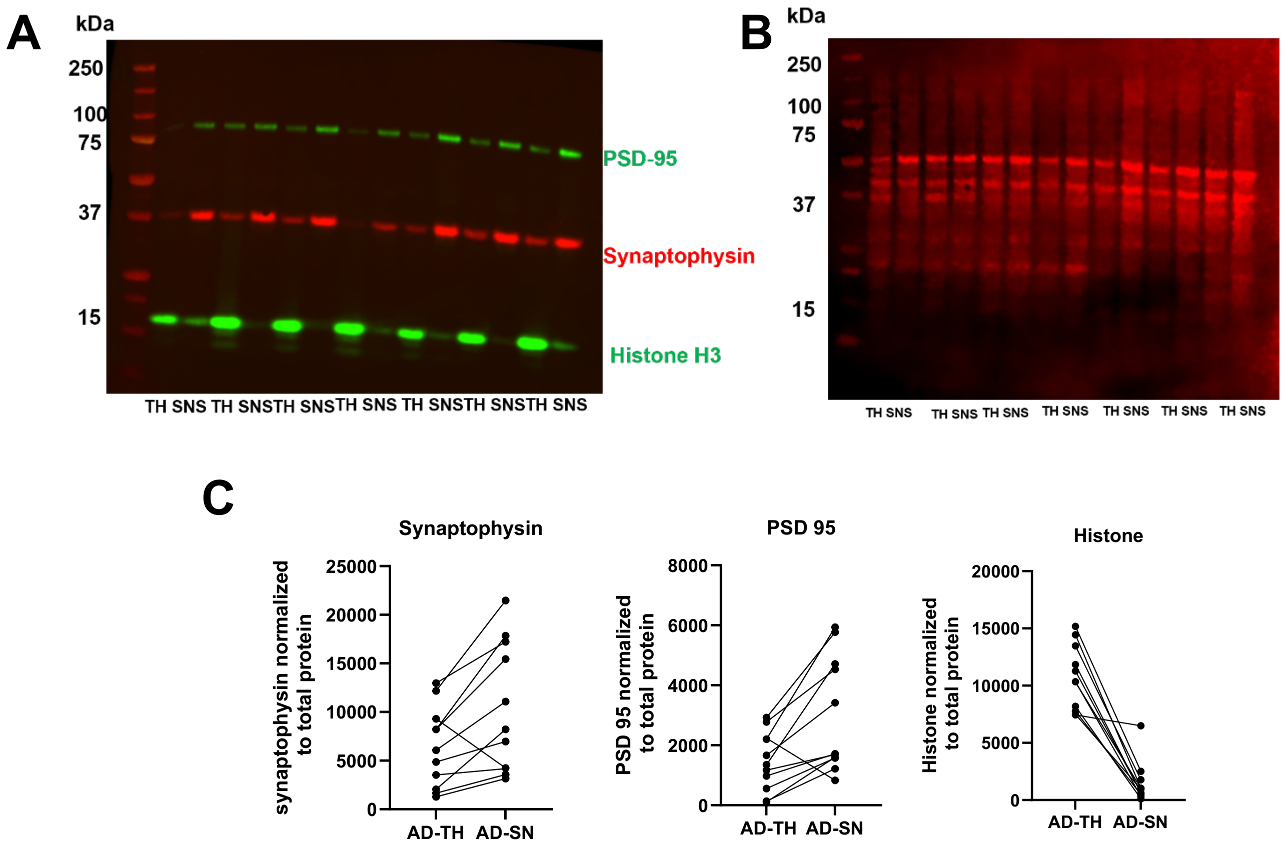


Wilcoxon matched-pairs signed rank test, n=11

PSD-95 p=0.0098; Histone p=0.001; Synaptophysin p=0.0186

**Supplementary figure 1:**

**Validation of synaptoneurosome and synaptosome preparations.** Image of a full-length Western blot (**A**), indicating whether a sample is from total homogenate (TH) or synaptoneurosomes (SN). Bands were quantified on Licor, and normalised to total protein stain shown in (**B**). Quantification (**C**) shows significantly increased protein levels of the pre- and post-synaptic markers synaptophysin and PSD-95, respectively, and decreased protein levels of histone (H3), indicating exclusion of non-synaptic material (Wilcoxon matched-pairs signed rank test).

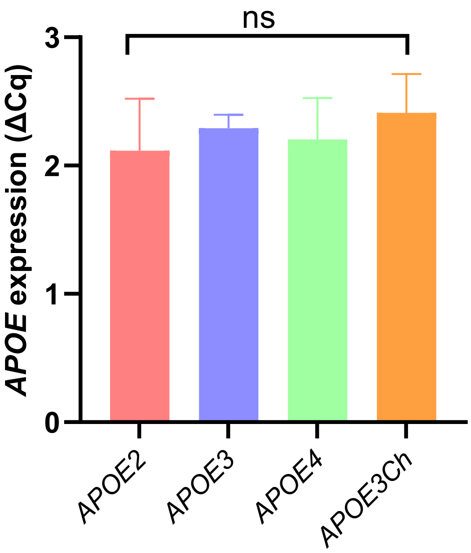


**Supplementary figure 2:**

**Comparable APOE expression across engineered cell lines normalised to actin**
qPCR analysis shows that APOE expression levels are consistent across all APOE-modified cell lines when normalised to the housekeeping gene actin, indicating that humanized APOE insertion does not alter expression levels.
